# A Systematic Review of the Etiology and Neurobiology of Intermittent Explosive Disorder

**DOI:** 10.1101/2024.09.12.24313573

**Authors:** John Paliakkara, Stacy Ellenberg, Andrew Ursino, Abigail A Smith, James Evans, Joseph Strayhorn, Stephen V. Faraone, Yanli Zhang-James

## Abstract

Intermittent Explosive Disorder (IED) is characterized by repeated inability to control aggressive impulses. Although the etiology and neurobiology of impulsive anger and impulse control disorders have been reviewed, no systematic review on these aspects has been published for IED specifically. We conducted a systematic search in seven electronic databases for publications about IED, screened by two authors, and retained twenty-four studies for the review. Our findings highlight a multifactorial etiology and neurobiology of IED, emphasizing the role of the amygdala and orbitofrontal cortex in emotional regulation and impulse control, and supporting interventions that target serotonergic signaling. Research also shows that childhood trauma and adverse family environment may significantly contribute to the development of IED. Yet, genetic studies focusing on IED were largely lacking, despite many examining the genetics underlying aggression as a general trait or other related disorders. Future research using consistently defined IED as a phenotype is required to better understand the etiology and underlying mechanisms and assist in informing the development of more effective interventions for IED.

## 1. INTRODUCTION

The Diagnosis of IED has evolved significantly over time within the Diagnostic and Statistical Manual of Mental Disorders (DSM) (Coccaro, 2000). Its closest original classification was described in DSM-I as a passive-aggressive personality (aggressive type) with irritability and temper tantrums, that later transformed into an explosive personality disorder in DSM-II (Association, 1952, 1968). In DSM-III, IED became its own diagnostic entity recognized as part of impulse control disorders (Association, 1980). It has undergone several revisions including IED Research criteria (IED-R), the DSM-IV criteria, and the IED Integrated Research criteria (IED-IR) (Association, 2000). IED-IR is a set of diagnostic criteria that aim to provide a more comprehensive definition of the condition by integrating the originally proposed research criteria (IED-R) with the DSM-IV criteria. The latest update, DSM-5, maintains the integrity of IED as a unique disorder, with the addition of A1 and A2 criteria mentioned earlier. Other related impulse control disorders that have been studied and reviewed alongside IED include kleptomania, pyromania, gambling disorder, and trichotillomania.

As defined in DSM 5, IED is characterized by impulsive aggression and can begin as early as age 6. It has a chronic course (Association, 2022; Coccaro et al., 2004; Kessler et al., 2006). These aggressive impulses can be triggered by minor events. The behavior is disproportionate to these perceived slights, and can’t be better explained by other medical, including psychiatric, conditions or an alternate organic cause. IED may lead to physical assault or property damage. It also can include non-physical acts of aggression such as verbal aggression. To meet diagnostic criteria, verbal aggression must occur on average twice weekly for 3 months, or physical aggression must occur at least 3 times within a 12-month period.

The person with IED often feels deep remorse for their actions afterward. Those diagnosed with IED feel intense emotions before their explosive rage episodes. These feelings are often accompanied by physiological symptoms such as tachycardia, diaphoresis, and dyspnea. After the explosion, they may feel a sense of relief and sometimes pleasure. However, most report feeling very upset, remorseful, embarrassed, and/or disappointed after their outbursts (Kulper et al., 2015).

In the U.S., life-time prevalence estimates for IED range from 5.4% to 7.3%; 12-month prevalence ranges from 2.7% to 3.9% (Kessler et al., 2006). IED has an early age of onset, ranging from 13.5 to 18.3 years of age (Coccaro et al., 2005; Coccaro et al., 2004; Kessler et al., 2006), and appears more common in males (Coccaro et al., 2005). IED is often comorbid with depression, anxiety, bipolar disorder, conduct disorder, oppositional defiant disorder, post-traumatic stress disorder, substance use disorder, antisocial personality disorder, and borderline personality disorder (Coccaro et al., 2005). Accurate assessment of IED can be difficult due to these comorbidities and the significant overlap of their features. The prevalence and psychiatric comorbidities may also vary across different races, underrepresented, or socioeconomically disadvantaged groups (Rees et al., 2013; Shevidi et al., 2023).

Despite many studies and reviews assessing the neurobiology of impulse control disorders and associated conditions characterized by impulsive anger, irritability or aggression, these valuable studies cover a wide range of trait expression but do not specifically address the IED diagnosis (Blair, 2016). No study has systematically explored both the neuroanatomy and neurophysiology of IED. Similar studies only examine these aspects in impulsive aggression. To address this gap, we undertook a systematic literature review aimed at delivering a comprehensive and critical examination of the existing research focused specifically on the etiology and neurobiology of IED. We sought to shed light on the underlying mechanisms of IED that would allow for a nuanced understanding of this disruptive disorder to inform clinicians and aid future research directions.

## 2. METHODS

### 2.1 Study Design

The systematic review was conducted in accordance with the Preferred Reporting Items for Systematic Reviews and Meta-Analyses (PRISMA) guidelines. The study protocol is registered in PROSPERO (registration ID CRD42024524230). The current review is one of the three separate studies within the registered protocol that examines various clinical aspects of IED. In accordance with our PROSPERO protocol, we used a diagnosis-focused, yet clinically broad search strategy to retrieve all publications that addressed IED as a specific diagnostic entity. Systematic search queries using the keyword phrase “intermittent explosive disorder” and associated controlled vocabulary (when available) were conducted in PubMed, EMBASE, CINAHL, Scopus, PsycINFO, Cochrane Library, and Web of Science collections. All databases were searched from inception to February 5, 2024. There were no search restrictions on language, publication status, or outcomes. The details of all the search strategies can be found in Appendix 1. Two independent reviewers screened the titles and abstracts of the identified records for relevancy. The full text of relevant papers was reviewed subsequently. Any disagreements between the reviewers were resolved by an independent third reviewer. The current report examines specifically the neurobiology (neuroanatomy and neurotransmitters) and etiology (psychosocial factors) of IED.

### 2.2 Inclusion and Exclusion Criteria

To be selected for the present review required that a paper examined participants of any age with a diagnosis of IED as defined by DSM III, III-R, IV, IV-TR, V, V-TR, IED-R, or IED-IR criteria. Studies that did not use the diagnosis of IED but examined related entities such as explosive aggression, or impulsive aggression, were not included. We focused our review on the diagnosis of IED to ensure it would be relevant to those diagnosed with IED specifically in clinical practice. Studies remained eligible for inclusion even if participants diagnosed with IED also fulfilled criteria for concurrent psychiatric or neurological disorders.

## 3. RESULTS

Twenty-four primary studies were identified that focused on neuroimaging, neurobiology, and psychosocial aspects of IED. Figure 1 shows the study selection process. The studies’ findings and other extracted variables can be found in Table S1. We describe our overall findings in three main areas: 1) structural, functional and connectivity differences in the amygdala, hippocampus, prefrontal cortex, orbitofrontal cortex, and hypothalamus 2) role of serotonin neurotransmitter system in IED 3) genetic and psychosocial factors for IED.

**Figure 1.**
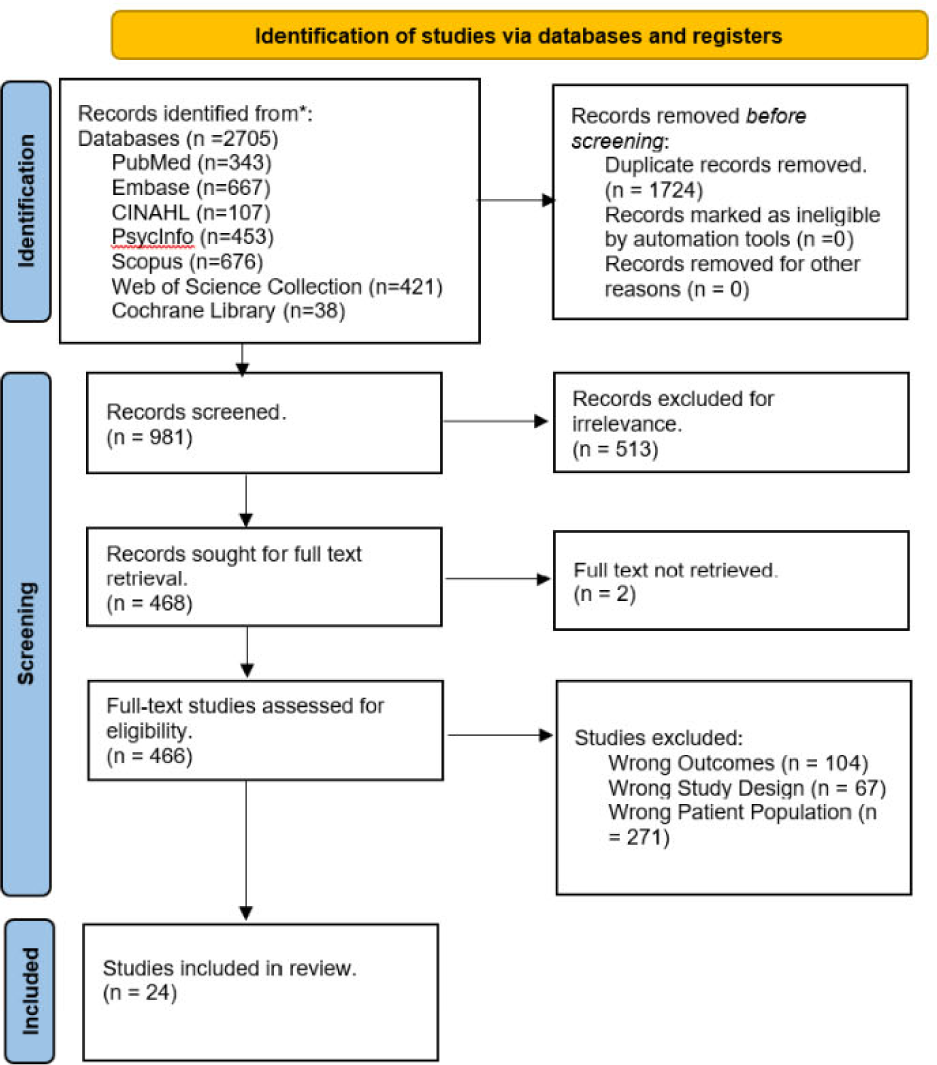
The PRISMA flow diagram for the systematic literature search, detailing the databases searched, the number of abstracts screened, the number of full texts screened, and the number of studies retained.

### 3.1 Neurobiology: Neuroimaging Studies

#### 3.1.1 Amygdala

The amygdala plays a crucial role in the processing of emotions, particularly fear and anger and in regulating impulsive behaviors and emotional responses (Coccaro, Keedy, et al., 2016; Coccaro et al., 2015; Coccaro et al., 2007; McCloskey et al., 2016).

People with IED scanned with a 3T MRI scanner exhibit heightened functional magnetic resonance imaging (fMRI) Blood Oxygen Level Dependent (BOLD) responses relative to healthy controls in their amygdala when exposed to social threats, such as angry faces (Coccaro et al., 2007; McCloskey et al., 2016). Additionally, an fMRI study measuring BOLD responses but with a 7T scanner has revealed that those with IED have more pronounced activation in the right amygdala during exposure to anger-inducing stimuli compared to controls (Seok & Cheong, 2020). Multiple studies also found from functional connectivity analyses that IED patients lacked functional connectivity between the amygdala and medial orbitofrontal cortex during processing of emotional stimuli relative to healthy controls. (Coccaro et al., 2007; McCloskey et al., 2016).

Compared with healthy controls and psychiatric controls, individuals with IED have smaller gray matter volumes in the right amygdala (Coccaro, Fitzgerald, et al., 2016). Conversely, Seok et al found that IED is associated with reduced gray matter volume in the left amygdala compared to controls (Seok & Cheong, 2020).

Another study found that while there is no difference between individuals with IED and healthy controls regarding the average size of the amygdala, there are significant differences in how the amygdala’s surface area changes in shape. Specifically, individuals with IED had inward deformations concentrated in the superior and inferior regions of the medial-anterior amygdala. The inward deformations could differentiate the two groups with 63% sensitivity, 71% specificity, and an area under the ROC of 0.724 (Coccaro et al., 2015), indicating that the amygdala differences were not seen in many IED patients.

#### 3.1.2 Frontal Cortex

The frontal cortex plays a crucial role in human aggression. It is a region of the brain that is involved in the regulation of emotions, decision-making, and behavior (Maley et al., 2010). Frontal cortex abnormalities have been found in some Individuals with IED that may contribute to their aggressive behavior. Abnormalities in the prefrontal cortex can lead to difficulties in inhibiting responses to provocation or stress, contributing to the aggressive outbursts characteristic of IED (Alvarez-Alonso et al., 2016).

Coccaro et al found that individuals with IED, compared with healthy controls and psychiatric controls, had a reduction in gray matter volume in orbitofrontal cortex, medial prefrontal cortex, anterior cingulate cortex (Coccaro, Fitzgerald, et al., 2016). Seok et al also found that IED was associated with reduced gray matter volume in the left orbitofrontal cortex compared to healthy controls (Seok & Cheong, 2020).

In an fMRI study, increased error-related activity was observed in the dorsolateral prefrontal cortex of individuals with IED. This was seen when performing tasks aimed at assessing impulse control, such as the color-word Stroop task (Moeller et al., 2014). Seok et al also found that individuals with IED had increased activation in the anterior cingulate cortex during exposure to anger-inducing stimuli compared to controls (Seok & Cheong, 2020).

The response to angry faces in the orbitofrontal cortex was weaker in individuals with IED compared to controls. Additionally, when viewing salient faces there were less pronounced responses in several regions of the prefrontal cortex among IED subjects compared to healthy controls (Coccaro et al., 2007; McCloskey et al., 2016). As mentioned earlier, McCloskey et al found decreased orbitofrontal cortex-amygdala functional connectivity in individuals with IED compared with healthy controls.

During fMRI scans, participants viewed video clips depicting social situations with either potentially aggressive acts or non-aggressive interactions. It was found that individuals with IED had less activation in the medial orbitofrontal cortex and anterior cingulate cortex than their healthy controls counterparts during the viewing of aggressive video clips. Additionally, there were reduced activations in the left ventrolateral prefrontal cortex as well as the bilateral inferior parietal lobules, precuneus, middle temporal gyri, and cerebellar hemispheres (Coccaro et al., 2022).

#### 3.1.3 Hypothalamus

The hypothalamus plays a crucial role in regulating hunger, thirst, sleep, and temperature regulation but is also involved in the control of emotions and behaviors, particularly those related to stress and aggression. The hypothalamus has been implicated in the modulation of this behavior due to its connections with other parts of the brain involved in emotion regulation, such as the prefrontal cortex and amygdala (Giordano et al., 2016).

The hypothalamus plays a critical role in the regulation of aggression in individuals with IED, particularly within the posterior region known as the posteromedial hypothalamus or "aggression area" (Giordano et al., 2016). This region is involved in generating sympathetic responses and is connected to various structures including the cingulate and insular cortices, bed nucleus of stria terminalis, and various amygdaloid nuclei through which it influences aggression and autonomic system activation (Contreras Lopez et al., 2021). Directional deep brain stimulation targeting this area can alleviate symptoms of refractory IED by allowing patients to control their aggressive behavior and suppress violent outbursts. This is supported by clinical reports where adjustments in deep brain stimulation settings have led to immediate calmer responses in patients, with long-term follow-up indicating sustained improvements in socialization, cognitive functions, and school performance (Contreras Lopez et al., 2021; Giordano et al., 2016; Maley et al., 2010).

The central role of the hypothalamus in IED pathogenesis suggests that targeting this area can be a viable therapeutic option for individuals with severe, drug-resistant aggressive disorders. Further research into the specific connections between the hypothalamus and other brain regions involved in IED would help to clarify its role in the disorder.

#### 3.1.4 Other Subcortical Structures

The surface morphology of the hippocampus is significantly different between individuals with IED and healthy controls. As mentioned earlier, the inward deformations of the amygdala found by Coccaro et al were able to be used to somewhat differentiate individuals with IED and healthy controls. The same study found similar inward deformations in the hippocampus. The inward deformations could differentiate the two groups with 61% sensitivity, 75% specificity, and a slightly lower area under the ROC of 0.715 (Coccaro et al., 2015).

Many other subcortical structures have been investigated for their role in aggression in patients with IED. These include the orbitofrontal projections along the inferior internal capsule and the ventral striatum. Deep brain stimulation for refractory IED has been seldom used in these regions with reported improvements in symptom severity and daily functioning in cases with refractory IED (Giordano et al., 2016; Maley et al., 2010).

In diffusion tensor imaging studies, individuals with IED have been found to have lower fractional anisotropy values in the superior longitudinal fasciculus, indicating potential disruptions in the white matter connections between frontal and temporoparietal regions (Lee et al., 2016).

Morphometric studies have found reductions in gray matter volume in the left insula and uncus as well. The severity of aggression in individuals with IED has been linked to lower gray matter volume in the insula (Coccaro, Fitzgerald, et al., 2016; Seok & Cheong, 2020). Functional MRI studies during anger-inducing stimuli have revealed increased activation in areas such as the putamen and left anterior insula in individuals with IED compared to healthy controls. This heightened activity may contribute to the intense emotional reactions seen in IED patients (Seok & Cheong, 2020).

In summary, the neurologic structures implicated in IED, including the hippocampus, and related subcortical regions, are critical for understanding the disorder’s underlying mechanisms and informing potential treatments like deep brain stimulation or pharmacological interventions aimed at modulating these brain regions.

### 3.2 Neurobiology: Role of Serotonin Systems in IED

While many studies of impulsive aggression or violence have documented the potential roles of catecholamine and glutamate neurotransmitter systems, our search on IED only identified studies that discussed serotonin. These studies have shown that individuals with IED often exhibit altered serotonin function compared to healthy controls. Reduced numbers of platelet 5-HT transporters are found in subjects with IED (Coccaro et al., 2010). Furthermore, ligand binding studies report alterations in 5-HT transporter availability in certain brain regions, such as the anterior cingulate cortex in IED patients with current physical aggression (Coccaro et al., 2010; Rosell et al., 2023).

Fluoxetine has shown anti-aggressive effects in IED patients and in subjects with a history of aggression, potentially due to its ability to increase synaptic serotonin levels and modulate neurotransmitter systems involved in aggression (Coccaro et al., 2009; Phan et al., 2011). However, the efficacy of fluoxetine varies among individuals with IED (Coccaro et al., 2009; Tahir et al., 2022). In one study examining the pharmacotherapy of IED, lorcaserin, a drug that targets the serotonin 2C receptor, was tested and found to have an effect on reducing provoked aggression during the Taylor aggression paradigm (Coccaro & Lee, 2019).

Rosell et al used positron emission tomography imaging to examine serotonin transporter availability (as measured by [11C] DASB binding) in brains of individuals with IED. It found that higher levels of trait aggression were linked to greater serotonin transporter availability in the anterior cingulate cortex and the ventral striatum. When adjusting for additional factors such as callousness traits and childhood trauma, these correlations became stronger. Additionally, baseline state aggression levels in IED patients were positively correlated with [11C] DASB binding in the anterior cingulate cortex. This research suggests that serotonin transporter availability may play a role in the expression of aggression related to IED (Rosell et al., 2023).

### 3.3 Etiology: Genetic and Psychosocial Risk Factors

#### 3.3.1 Family Studies

While studies examining the genetic basis for IED are limited, clinical observations and family history data suggest that IED is a heritable condition. Coccaro et al investigated this utilizing the Integrated Research Criteria for Intermittent Explosive Disorder (IED-IR). Their study has demonstrated that there is a significant difference in the morbid risk of developing IED-IR among first-degree relatives of patients with IED-IR compared to those of control subjects without the disorder (p < 0.00001) (Coccaro, 2010). The increased morbid risk for IED was not influenced by comorbid conditions (specifically borderline personality disorder and/or antisocial personality disorder) among the probands or their relatives (p < 0.0005).

#### 3.3.2 Early Life Experiences and Family Environment

The research into the relationship between early life experiences, family environment, and IED suggests that there is a significant link between adverse childhood events, such as trauma, abuse, and neglect, and the later development of IED. The Parenting Behavior Inventory (PBI) is a self-rating scale measuring the qualitative aspects of parental behavior. The PBI has two subscales: Parental Care and Parental Control. The Parental Care (PBI Care) subscale assesses perceived care, including affection, emotional warmth, empathy, and closeness versus emotional coldness, indifference, and neglect. The Parental Control subscale measures intrusiveness and parental direction through guilt or encouragement of autonomy and independent thinking. PBI has been used to assess self-reported parental care among individuals with IED, revealing lower scores for care compared to both psychiatric controls and healthy controls, which suggests a lack of supportive family environment during childhood. Additionally, higher PBI Control scores among IED subjects and psychiatric controls indicate that the parenting style may have been more controlling, or authoritarian compared to parenting styles used for healthy controls, potentially leading to feelings of oppression, and possibly contributing to aggression in these individuals (Lee et al., 2014).

The logistic regression analyses further confirm an association between IED and a range of interpersonal traumatic events experienced during childhood (Nickerson et al., 2012; Puhalla et al., 2020). There were multiple variables found to be statistically significant predictors of an IED diagnosis. These variables were analyzed in the context of first exposure in childhood and adulthood. The variables with odds ratios statistically significant only in first exposure in childhood included badly beaten by parents, raped, sexually assaulted, witnessed serious physical fights at home as a child, and saw someone badly injured/killed. The variables with significance in both childhood and adulthood included badly beaten by partner, badly beaten by someone else, and mugged. While these three variables were significant in both childhood/adulthood first exposure, the childhood odds ratios were significantly larger than the adulthood odds ratios. This difference was supported by Chi-Square analyses with p values < 0.001 demonstrating the statistically greater impact on IED diagnosis these variables had when exposed in childhood. These findings highlight the importance of early life experiences on the development of psychological disorders such as IED. Shevidi et al found that participants with IED reported more extensive abuse and neglect histories compared to healthy individuals and those with non-IED psychiatric disorders, reinforcing the link between trauma exposure and violent behavior associated with IED (Shevidi et al., 2023).

#### 3.3.3 Psychosocial factors

As mentioned earlier, socioeconomic status data from multiple studies reviewed here show mixed results of negative or no correlation with IED diagnosis. Shevidi et al found that their sample of 478 individuals with IED had an average lower socioeconomic status score compared to their 215 psychiatric controls and 452 healthy controls. The average socioeconomic status was 38.9 for IED, 42.3 for psychiatric controls, and 47.0 for healthy controls. Their statistical analyses showed significant differences between each group (IED < psychiatric controls < healthy controls) and this trend has been found in other studies as well (Lee et al., 2014; Shevidi et al., 2023). However, there are also other studies with similar population sizes that report no statistically significant differences in socioeconomic status (Fanning et al., 2014). Other studies reviewed here also report socioeconomic status scores with mixed results regarding statistically significant differences between groups, however those were neuroanatomical and neurobiological studies that understandably had much lower population sizes (Coccaro et al., 2022; Coccaro et al., 2015; Coccaro et al., 2007).

Socioeconomic status has been further investigated due to its role as a risk factor for exposure to trauma, with those from lower socioeconomic backgrounds being more likely to experience traumatic events. In Timor-Leste, where the country has faced periods of peace and internal conflict and has had ongoing socioeconomic challenges since its establishment in 2002, these factors have been linked to the prevalence of IED among the population (Rees et al., 2013). The research by Rees and colleagues indicates that IED is associated with a history of trauma exposure, particularly from conflict-related events. Furthermore, they suggest that the socioeconomic environment plays a crucial role in maintaining these anger attacks even after conflicts have ceased, due to persistent challenges such as high unemployment, poverty, and limited access to healthcare. The specific trauma variables investigated for their impact on IED diagnosis included “Suffered because helped resistance”, “Suffered serious trauma as a woman” (gender-based violence such as rape), “Severe poverty index”, “Family and Community conflict index”, and “Distressing preoccupations with injustice”. All these variables, except gender-based violence, showed a positive correlation with IED diagnosis. As mentioned earlier, Nickerson et al found that rape and sexual assault in childhood, but not adulthood, was statistically significant in predicting IED diagnosis, but the study by Rees et al only surveyed adults. The other variables demonstrated higher prevalence in individuals with IED and were shown to be statistically significant predictors: “Suffered because helped resistance” (Suffered: 31.8% IED vs Non- Suffered: 8.5% IED, OR=2.33[1.48, 3.68]), “Severe poverty index” (IED: Mean=2.45[SD=1.88] vs Non-IED: Mean=1.80[SD=1.64], OR=1.23[1.08, 1.39]), “Family and Community conflict index” (IED: Mean=0.91[SD=0.93] vs Non-IED: Mean=0.52[SD=0.82], OR=1.88[1.27, 2.77]), “Distressing preoccupations with injustice” (None: 7.1% vs ≥2 historical periods: 23.3%, OR=2.10[1.35, 3.28]). Post conflict distress also mediates the relationship between trauma exposure and socioeconomic disadvantage, which further underscores the impact of socioeconomic status on the development and maintenance of IED symptoms.

## 4. DISCUSSION

The diagnostic concept of IED has undergone significant changes over the past several decades, reflecting a refined understanding of clinical symptomatology and a drive to improve diagnostic accuracy and differentiation from other disorders with similar symptoms. As the first systematic review focused on the neurobiology and etiology of IED through a targeted approach, our study offered a comprehensive overview of the current state of research on IED. It illuminated several crucial aspects of the etiology and neurobiology of IED. These aspects are the foundations of diagnosis, assessment, and treatment. By studying a clearly defined DSM-diagnosis, consistent research can be reproduced. Diagnostic criteria and treatment protocols can be properly advanced as researchers continue to measure outcomes. Research that uses inconsistently defined terms such as “impulsive aggression” can’t be translated as effectively into clinical guidelines, as how one defines “impulsive aggression” varies from study to study. These inconsistencies in definitions may trickle into the quality of evidence seen in primary studies as well if the population characteristics are not clearly defined by structured diagnostic criteria.

Our review found that most of the literature in the specific context of IED were neuroimaging studies. These studies provide evidence for supporting the involvement of various brain regions, including the amygdala, hippocampus, prefrontal cortex, orbitofrontal cortex, and hypothalamus. fMRI studies indicate that individuals with IED exhibit altered BOLD responses in these areas when exposed to social threats or anger-inducing stimuli compared to controls. Additionally, reduced gray matter volume has been observed in IED patients, which may contribute to difficulties in regulating impulsive behaviors. Pharmacological interventions targeting serotonergic pathways show some efficacy in managing aggressive symptoms, suggesting a role for serotonin in IED pathology. Studies on psychosocial factors supported the significant roles that early life experiences and family environment, for example, adverse childhood events and controlling or authoritarian parenting styles, may have in the development of IED. Socioeconomic status is another risk factor, as individuals from lower socioeconomic backgrounds are more likely to experience trauma, which can contribute to the development and maintenance of IED symptoms, particularly in post-conflict settings. Although there are very few genetic studies of IED, a family study suggests that it may be a heritable condition, like other disorders with aggressive symptoms.

### 4.1 Neurobiology

Functional MRI studies suggest people with IED exhibit increased amygdala activation to social threats and structural brain anomalies, including reduced functional connectivity between the amygdala and medial orbitofrontal cortex and altered gray matter volume in specific regions of the amygdala. Deep brain stimulation in the posteromedial hypothalamus improves symptoms of refractory IED, including immediate calmer responses and long-term improvements in socialization, cognitive functions, and occupational performance. The hippocampus shows altered surface morphology in individuals with IED, and lower fractional anisotropy values in the superior longitudinal fasciculus suggest disrupted white matter connections affecting processing of social and emotional information. Functional MRI studies during anger-inducing stimuli have demonstrated increased activation in other subcortical structures like the putamen and anterior insula, which may contribute to the intense emotional reactions that are characteristic of IED.

These findings collectively suggest that a combination of abnormalities across multiple brain regions, including those of the limbic system, are implicated in IED pathology and could inform targeted treatments for the disorder. Due to the search strategy, literature that did not assess IED status, but instead studied the role of the amygdala and cortical areas in impulsive aggression were not included. However, they did reach similar interpretations of their findings relative to the studies reviewed here (da Cunha-Bang et al., 2019; Siep et al., 2019; Soloff et al., 2017).

Serotonin function appears to be altered in individuals with IED, as indicated by reduced serotonin transporter availability and alterations in receptor activity, which may contribute to the expression of aggression. Fluoxetine has been shown to have anti-aggressive effects in IED patients, however, individual responses to fluoxetine vary. Additionally, the increased availability of 5-HT2A receptors in the orbitofrontal cortex has been correlated with state measures of impulsive aggression, suggesting a role for these receptors in controlling aggressive behavior. Before informing clinical practice, replication of the fluoxetine data is needed given many studies that report unintended and contradictory effects of SSRIs, where treatment groups had worsened rates of suicidality, aggression, and violent crime (Lagerberg et al., 2020; Molero et al., 2015; Pappadopulos et al., 2006; Sharma et al., 2016).

Other neurotransmitter systems may also play a significant role in IED. Studies have implicated imbalances in dopamine signaling (levels or receptor activities) that may lead to frustration and impulsive aggression (Retz et al., 2003; Schluter et al., 2013). Certain antipsychotic drugs that modulate dopamine activity can also reduce aggressive behavior, supporting the link between dopamine and aggression. Genetic factors such as the DRD2 Taq A1 and DAT1 polymorphisms have been linked to an increased risk for impulsive aggression (Modestino et al., 2022). A study of 95 human Mendelian disorders that documented symptoms of aggressive behaviors found that their causal genes were significantly enriched in both dopamine and serotonin signaling pathways (Zhang-James & Faraone, 2016). Glutamate levels are positively correlated with aggression and impulsivity, supporting its role in promoting these behaviors through excitatory neural circuitry (Fanning et al., 2020; Lee & Coccaro, 2019). Many other studies assess other neurotransmitters; however, they focus on impulsivity or aggression as opposed to a DSM-defined disorder (Chester et al., 2016; Ende et al., 2016; Retz et al., 2003; Schluter et al., 2013). Our search strategy, however, was unable to find studies that investigated other neurotransmitter systems except serotonin systems. Future studies are needed to assess other neurotransmitter systems such as dopamine, norepinephrine, and glutamate, as well as their interactions with serotonin signaling, in the underlying pathophysiology of IED.

### 4.2 Etiology

While genetic studies are limited, family history data suggest that IED may have a heritable component, evident from statistically significant increased morbid risk in first-degree relatives of individuals with IED, that remained significant when accounting for common comorbidities. While our search was unable to find candidate gene studies and genome-wide association studies with populations diagnosed with IED, there is a large volume of genetic literature investigating the genetic basis of aggression as a trait or other disorders with aggression (Demontis et al., 2021; Veroude et al., 2016; Zhang-James & Faraone, 2016; Zhang-James et al., 2019). Earlier twin and adoption studies generally estimated that about 50% of the variance in broadly defined aggressive behavior in humans can be explained by genetic influence (Tuvblad & Baker, 2011; Waltes et al., 2016). Recent genome-wide association studies (GWAS) have also often focused on more broadly defined, or related disorders such as disruptive behavior disorders, conduct disorder, and antisocial behavior (Demontis et al., 2021; Musalkova et al., 2024; Rose et al., 2004; Slutske et al., 1997; Tielbeek et al., 2022). While a multi-trait or transdiagnostic approach is often needed to enhance the statistical power, as demonstrated by their success of the discovery of genome-wide significant loci and findings of significant genetic correlations between aggression phenotype and other psychiatric disorders, the translation of these genetic findings to the etiology of IED specifically still poses an additional challenge (Demontis et al., 2021; Tielbeek et al., 2022; Waszczuk et al., 2023).

A complex interplay exists between early life experiences, family environment, and the development of IED. From Lee et al, a lack of supportive family environment during childhood and a more controlling/authoritarian parenting style was observed among IED individuals. Several studies reported an association between IED, and various interpersonal traumatic events experienced during childhood. Additionally, participants with IED reported more extensive histories of abuse and neglect than healthy controls and psychiatric controls. These reinforce the connection between childhood trauma exposure associated with IED.

Socioeconomic status emerges as another critical risk factor for exposure to trauma, with lower socioeconomic backgrounds being more prone to traumatic events. In Timor-Leste, where the country has faced conflict and ongoing socioeconomic challenges, IED prevalence is linked to a history of trauma exposure and lack of resources.

Gene and environmental interactions have also been supported by twin and adoption studies as well as GWAS studies of broadly defined aggressive behavior or related disorders (Rose et al., 2004; Slutske et al., 1997). These disorders demonstrated significant genetic correlation with traits such as educational attainment and age at first birth, suggesting an interaction with psychosocial and environmental factors. Polygenic risk for aggression was found to be associated with exposure to traumatic events (Goodman et al., 2004; Magwai & Xulu, 2022; Sadeh et al., 2016). Additionally, epigenetic studies suggest that non-genetic risk factors can also influence the stress response and the development of aggressive behaviors (Waltes et al., 2016). These findings of the interaction between genetic susceptibility and environmental factors emphasize the complex multi-factorial etiology of aggressive behavior. Nevertheless, future studies are needed to further identify and clarify the underlying genetic and environmental mechanisms of IED specifically,

### 4.3 Future Research and Limitations

The primary limitation of this study stems from the nature of our highly specific search strategy focusing on the diagnosis of IED, excluding studies only discussing impulsive aggression and related traits. While our search strategy was beneficial in isolating studies of IED, there are other studies of samples that potentially met IED criteria but were not formally given a diagnosis of IED. Another limitation is that many studies found through the systematic search studied populations with IED and various comorbidities. A significant proportion of the work done in studying the etiology/neurobiology of IED and improving diagnosis, assessment, and treatment, has been completed by a smaller number of research groups. This may create bias in how IED is studied if samples are selected from nearby communities. Another limitation was the heterogeneity of study methods in analyzing the various aspects of IED. While this review has many limitations, studies independent from these groups, such as this review, are important in supporting the validity of the research already done.

While these are significant limitations, they also highlight a potential underlying issue of the lack of focus on IED in research studies of impulsive aggression. When studies do not use standardized criteria that can be referenced to clinical practice, their results become more difficult to translate to the clinical context.

## 5. CONCLUSION

As the first systematic review of the neurobiology and etiology of IED, our findings implicate the amygdala, hippocampus, prefrontal cortex, orbitofrontal cortex, and hypothalamus as key to the neurobiology of IED, and underscore the significant role of serotonin signaling. Additionally, our findings also highlight the impact of adverse childhood experiences, family environment, and socioeconomic status in the development of IED. Although IED may be a heritable disorder, the genetic basis of IED is not yet fully studied. Future studies with populations of clearly diagnosed IED are needed to help improve the understanding of the neurobiology and etiology of IED specifically, but also clarify the differences from and relationship to other related disorders.

### Declaration of Interest

SVF received income, potential income, travel expenses continuing education support and/or research support from Aardvark, Aardwolf, AIMH, Akili, Atentiv, Axsome, Genomind, Ironshore, Johnson & Johnson/Kenvue, Kanjo, KemPharm/Corium, Noven, Otsuka, Sky Therapeutics, Sandoz, Supernus, Tris, and Vallon. With his institution, he has US patent US20130217707 A1 for the use of sodium-hydrogen exchange inhibitors in the treatment of ADHD. He also receives royalties from books published by Guilford Press: *Straight Talk about Your Child’s Mental Health*, Oxford University Press: *Schizophrenia: The Facts* and Elsevier: ADHD: *Non-Pharmacologic Interventions.* He is Program Director of www.ADHDEvidence.org and www.ADHDinAdults.com. Dr. Faraone’s research is supported by the European Union’s Horizon 2020 research and innovation programme under grant agreement 965381; NIH/NIMH grants U01AR076092-01A1, R0MH116037, 5R01AG064955-02, 1R21MH126494-01, 1R01NS128535-01, R01MH131685-01, 1R01MH130899-01A1, Corium Pharmaceuticals, Tris Pharmaceuticals and Supernus Pharmaceutical Company. JP, SE, AU, JS, and YZJ have no disclosures to report.

## Data Availability

All data produced in the present work are contained in the manuscript

## FUNDING

This research did not receive any specific grant from funding agencies in the public, commercial, or not-for-profit sectors.

## APPENDIX 1

### PubMed

"intermittent explosive disorder*"[Title/Abstract]

### Embase

**#1** ‘intermittent explosive disorder’/exp OR ‘intermittent explosive disorder*’:ti,ab

### CINAHL

**S1** TI ("intermittent explosive disorder*" OR AB "intermittent explosive disorder*")

### Scopus

TITLE-ABS-KEY ("intermittent explosive disorder*")

PsycInfo:

**S1** MAINSUBJECT.EXACT ("explosive disorder") OR tiab ("intermittent explosive disorder*")

### Cochrane Library (Cochrane Database of Systematic Reviews; CENTRAL)

**#1** intermittent NEXT explosive NEXT disorder*

### Web of Science Collections ((SCI-EXPANDED; ESCI; CPCI-S; SSCI)

#1 TS=("intermittent explosive disorder*")

